# Transcriptome-wide analyses delineate the genetic architecture of expression variation in atopic dermatitis

**DOI:** 10.1101/2024.11.21.24317734

**Authors:** Charalabos Antonatos, Dimitra Mitsoudi, Alexandros Pontikas, Adam Akritidis, Panagiotis Xiropotamos, Georgios K. Georgakilas, Sophia Georgiou, Aikaterini Tsiogka, Stamatis Gregoriou, Katerina Grafanaki, Yiannis Vasilopoulos

## Abstract

Genome-wide association studies (GWASs) for atopic dermatitis (AD) have uncovered 81 risk loci in European participants, however translating these findings into functional and therapeutic insights remains challenging. We conducted a transcriptome-wide association study (TWAS) in AD leveraging *cis*-eQTL data from 3 central AD tissues and the latest GWAS of AD in Europeans. We implemented the OTTERS pipeline that combines polygenic risk score (PRS) techniques accommodating diverse assumptions in the architecture of gene regulation. We also used differential expression datasets and co-expression networks to characterize the transcriptomic landscape of AD. We identified 176 gene-tissue associations covering 126 unique genes (53 novel). Most TWAS risk genes were identified by adaptive PRS frameworks, with non-significant differences compared to clumping and thresholding approaches. The novel TWAS risk genes were enriched in allergic reactions (e.g., *AQP7*, *AFF4*), skin barrier integrity (e.g., *ACER3*) and inflammatory pathways (e.g., *TAPBPL*). By integrating co-expression networks of lesional AD skin, we identified 16 hub genes previously identified as TWAS risk genes (6 novel) that orchestrate inflammatory responses (e.g., *HSPA4*) and keratinization (e.g., *LCE3E*, *LCE3D*), serving as potential drug targets through drug-gene interactions. Collectively, our findings provide additional risk genes for AD with potential implications in therapeutic approaches.

## Introduction

Atopic dermatitis (AD) was historically considered an allergic reaction and the precursor of the atopic march^1^. AD is now understood to involve complex interactions between skin barrier deficiency, systemic T helper (T_H_)-2 skewed inflammation and an increasing role of Th1 and Th17 inflammatory processes in chronic phases^2^. AD displays a strong genetic background, with heritability estimates reaching up to 80% from twin studies^3^. Large-scale genome-wide association studies (GWASs) uncovered 81 loci enriched in various inflammatory pathways apart from skin barrier integrity^4^ However, the genetic architecture of AD remains only partially resolved. GWASs often identify lead variants within broad haplotype blocks, which complicates pinpointing specific functional mechanisms. Moreover, many AD-associated loci map to regulatory elements, as is the case with most polygenic disorders^5^, thus restricting the direct translation of genetic findings into biological pathways that may serve as therapeutic targets.

The above non-coding variants are postulated to affect complex traits by modulating additional layers of -omics information, including gene expression. This concept has driven considerable efforts into mapping expression quantitative trait loci (eQTLs) and their combination with genetic evidence^6^. Transcriptome-wide association studies (TWASs) exploit tissue-specific expression reference panels to construct per-gene expression prediction models according to *cis*-eQTL variants (that is, variants withing 1Mb of transcription start and end sites)^7^. These models are then used to impute genetically regulated gene expression. Recent advances now permit the use of summary-level *cis*-eQTL and GWAS data for predicting gene expression and subsequently performing gene-based association tests. Compared to traditional GWAS, TWASs enhance interpretability, prioritize likely causal genes for disease risk, and improve statistical power, leading to widespread application across a range of complex diseases^8^.

Previous application of TWASs that leverage summary-based *cis*-eQTLs and GWASs have been applied in AD, including summary-based mendelian randomization (SMR)^9^, FUSION^10^ and MR-JTI^11^, leading to significant improvements in understanding AD pathophysiology. To prioritize AD-relevant genes, Sobczyk et al. performed an exhaustive gene-based prioritization using Genotype-Tissue Expression (GTEx) V7^12^ data among others, revealing a major inflammatory network that drives AD onset, complementary to genes that modulate skin barrier integrity^13^. On the contrary, Song and colleagues attempted to highlight novel drug targets through TWASs and GTEx v7 expression datasets, showcasing several therapeutic approaches that are relevant for skin conditions and reported a modest structural similarity with approved AD pharmacotherapies^14^. Wu et al. conducted a joint-tissue prioritization of candidate genes for AD, combining evidence from GTEx v8^15^ datasets and large-scale GWASs from meta-analysis of large-scale biobanks and consortia to identify 7 novel risk genes for AD, including *AQP3* and *PDCD1*^16^.However, each TWAS method employed has specific assumptions about the genetic architecture underlying gene expression, potentially leading to missed associations. Additionally, recent expansions in genome-wide analyses for AD have almost tripled the number of identified risk loci^17^, thereby facilitating the identification of novel AD risk genes.

In this study, we triangulated transcriptomic data to identify novel risk genes for AD and to dissect the genetic architecture underlying expression variation in cutaneous inflammation. We leveraged large-scale genome-wide data in AD, blood and skin *cis*-eQTLs, and implemented a novel approach combining PRS techniques that accommodate diverse assumptions in the architecture of gene regulation. Additionally, we incorporated RNA sequencing (RNA-seq) data to elucidate further pathophysiological links and co-expression networks not accessible through genetically regulated gene expression alone. This functional transcriptomic landscape provides a framework for characterizing the genetic basis of expression regulation in AD and guiding future therapeutic development through biologically-informed insights.

## Materials and Methods

### Data Sources

We leveraged the largest GWAS for AD to date, comprised of >800000 participants of European ancestry^17^. Three *cis*-eQTL summary statistics for 3 AD-relevant tissues, including whole blood (WB; n=670), sun exposed skin (SE; n=602) and non-sun exposed skin (NSE; n=517) from the GTEx v8 study^12^. Although tissue enrichment analyses have revealed a broader range of AD-related tissues^18^, we restricted our study to the above 3 to focus on those most representative of systemic and localized AD pathophysiology. GWAS data for AD were converted from the GRCh37/hg19 to the GRCh38/hg38 version using the UCSC liftOver tool, excluding variants with a minor allele frequency (MAF)>0.01. Linkage disequilibrium (LD) computations relied on an external reference of 503 European unrelated samples from the from the 1000 Genomes project^19^.

We systematically searched the Gene Expression Omnibus (GEO) database through December 4, 2023, for total RNA-seq datasets in adult human samples. For eligible AD datasets, we restricted analyses to lesional skin against healthy controls. We focused on skin samples, as the site of inflammation (that is, central tissue) is known to accurately represent the molecular pathomechanisms of a disease compared to peripheral tissues^20^.

### Omnibus transcriptome-wide association study

We conducted TWAS using OTTERS, an omnibus transcriptome test combining polygenic risk score (PRS) models with *cis*-eQTL training in 2 distinct stages^21^. OTTERS enhances statistical power by integrating multiple PRS models with different assumptions about the *a priori* unknown genetic architecture of polygenic traits. In specific, OTTERS implements 5 distinct methods for PRS calculations: (i) P-value thresholding of P-value<0.05 and (ii) P-value<0.001 with LD clumping (P+T)^22^, (iii) elastic net regression through lassosum^23^, (iv) Bayesian regression with a continuous shrinkage (PRS-CS)^24^, and (v) a non-parametric Bayesian multiple Dirichlet progress regression method (SDPR)^25^. The PRS-CS method was omitted to reduce computational burden as per authors’ recommendations.

At Stage I, OTTERS derives *cis*-eQTL weights per training method and performs a gene-based association analysis in the GWAS dataset, producing respective Z-scores and P-values. At Stage II, OTTERS applies an aggregated Cauchy association test (ACAT)^26^ to generate an omnibus P-value (ACAT-O). ACAT-O P-values were adjusted for genomic control in a tissue-specific manner to ensure that the median observed OTTERS p-value aligned to the 0.5 expected value under the null hypothesis^27^. We declared significant associations if (i) the ACAT-O P-values passed the Bonferroni-corrected P-value threshold at each tissue, (ii) the Z-scores produced were concordant among all PRS methods at each tissue, and (iii) at least 2 PRS methods produced nominally significant results (P-value_tissue_<0.05) at each tissue. Z-scores yielding NA in any PRS method were set to 0. Differences in the number of identified genes between each method were calculated with chi-squared tests. Functional enrichment analysis for each tissue was performed in the Reactome database through a hypergeometric test, with significant terms shown by a false discovery rate (FDR)<0.05. We declared TWAS risk genes as novel if they have not been previously reported in the TWAS atlas database^28^ and the latest published TWAS in AD to date^16^.

### Colocalization of TWAS risk genes

We applied Bayesian colocalization under a single causal variant hypothesis using the coloc v5.2.3R package^29^. Colocalization analysis computes the posterior probability (PP) of 5 different hypotheses, with PP.H3 indicating separate causal variants and PP.H4 indicating a shared causal variant. A threshold of PP.H4>0.5 was adopted to declare strong evidence for colocalization in TWAS risk genes across all 3 tissues as performed previously^13^.

### RNA-sequencing analyses

Publicly available RNA-seq data were re-analyzed using aPEAch, a tool for next generation sequencing pre-processing and mapping^29^. We performed quality assessment of raw reads with FastQC v0.12.1, as well as removed adapter sequences and low-quality reads through TrimGalore v0.6.10 with parameters “-q 20 –stringency 3”. We then aligned RNA-seq reads with STAR v2.7.10^30^ to the GRCh38/hg38 reference genome assembly (Ensembl version 107) restricting multimapping, removed duplicated reads with Picard Tools v3.0.0 and filtered mitochondrial reads with Samtools v1.17^31^. The count matrix was generated using featureCounts (SubRead package, v2.0.3)^32^ with the parameters “-p” for paired-end data whenever applicable and “-s” for strand specificity as determined through the RSeQC package^33^. Study-specific count matrices were normalized using the limma package^34^ and submitted to DExMA for random-effects meta-analysis^35^. We chose DExMA due to the build-in imputation of missing gene expression by applying the k-nearest neighbors (KNN) in the space of samples method^36^, an expected inter-study divergence. We declared significant results according to FDR≤0.01 and log_2_|Fold Change|(log_2_|FC|)≥1. Between-study heterogeneity was visualized with Quantile-Quantile (QQ) plots. Summary results underwent gene-set enrichment analysis (GSEA) using the KEGG pathways database. To assess whether the meta-analysis signature was associated with drug responses, we retrieved meta-analyzed data from the connectivity map (CMAP) build 02 drug signatures and small molecule perturbagens through the LINCS L1000 dataset^37^. Next, we calculated the cosine similarity between the meta-analysis transcriptome profile and the intersection of CMAP build 02 and LINCS L1000 datasets. Computations were performed with the ccmap Bioconductor R package^38^.

We also performed a weighted gene co-expression network analysis (WGCNA) to identify potential transcriptional interactions between TWAS-prioritized genes and dysregulated patterns of AD inflammation^39^. To reduce the inherent heterogeneity in high-throughput experiments, we allowed only paired-end data in the WGCNA approach, as performed elsewhere^40^. Combined paired-end datasets were filtered for low-count transcripts, retaining the top 75% genes according to their variance, corrected for batch effects and normalized with variance-stabilizing transformation^41^. Outliers were detected and excluded with Hubert’s algorithm^42^. The batch-corrected, normalized count matrix was submitted to principal component analysis (PCA) for visual exploration and WGCNA. As a rule of thumb, the soft thresholding power was optimized to balance high scale-free topology (R^2^>0.8) and mean connectivity below 100. We calculated network adjacencies, transformed to scaled signed topological overlap matrices (TOMs), and hierarchically clustered to identify the corresponding modules. The deepSplit parameter was set at 4, mergeCutHeight at 0.2 and the minModuleSize at 20. Correlation patterns between each eigengene expression and disease status were computed via Pearson correlation. Module preservation statistics were computed to estimate the robustness of identified co-expression networks using the same set as both reference and test sets. Hub genes were defined with module membership (MM)>0.8 and gene significance (GS)>0.2. Overlapping hub genes with TWAS risk genes were analyzed in DGIdb v5.0 to identify potential drug-gene interactions^43^.

## Results

### Transcriptome-wide association results across all tissues

Through the OTTERS framework, we obtained summary-level Z-scores and P-values for each of the 4 discrete PRS models in each tissue. We observed moderate inflation in the ACAT-O P-values through QQ plots and adjusted each method in each tissue for genomic control (Figs. S1-S3). By further applying Z-score and nominal P-value criteria (Methods), we identified 47 significant genes in WB (Table S1), 65 in SE (Table S2) and 64 in NSE (Table S3) at a tissue-specific Bonferroni-corrected P-value threshold (Fig. 1a). Several genes were detected in >2 tissues, including well-known AD risk loci such as *RPS26*, *SUOX*, *FLG* and *KIF3A* among others. We next evaluated whether TWAS signals were driven by proximal associations with variants passing the genome-wide significance (GWS) threshold (P-value<5×10^-8^). We assessed the overlap of 1Mb window around each TWAS risk signal with a 1Mb window around each GWS variant. In total, we identified 10 unique genes (9 novel), including *CFAP96*, *ZBTB5*, *ACADSB*, *XRRA1* (shared between WB and SE), *NCAPD2*, *TAPBPL* (shared between WB and SE), *IGFLR1*, *ZMYND8*, *GATD3* (shared between SE and NSE) and *PWP2* (shared between SE and NSE). Compared to previously reported TWAS risk genes in the TWAS atlas^28^ and the latest TWAS in AD^16^, we identified 53 unique novel TWAS risk genes (Fig. 1a). Some of these genes are implicated in biological activities relevant for AD. For example, the aquaglyceroporin *AQP7* is expressed in Langerhans cells modulating allergy induction^44^ and showed overexpression in SE and NSE (Fig. 1a). Similarly, the *AFF4* transcription elongation factor within the 5q31.1 locus^45^ has been associated with increased asthma risk through genetically predicted overexpression in lung^46^; *AFF4* further displayed a concordant sign in our study across all 3 tissues (Fig. 1a). *ACER3* is an alkaline ceramidase that participates in sphingolipid metabolism, a vital pathway in formation of the skin barrier interacting with ceramides and structural keratinocyte proteins significantly associated in our TWAS (e.g., *FLG*, *SPRR1B*, *SPRR2B*)^47^.

**Fig. 1.**
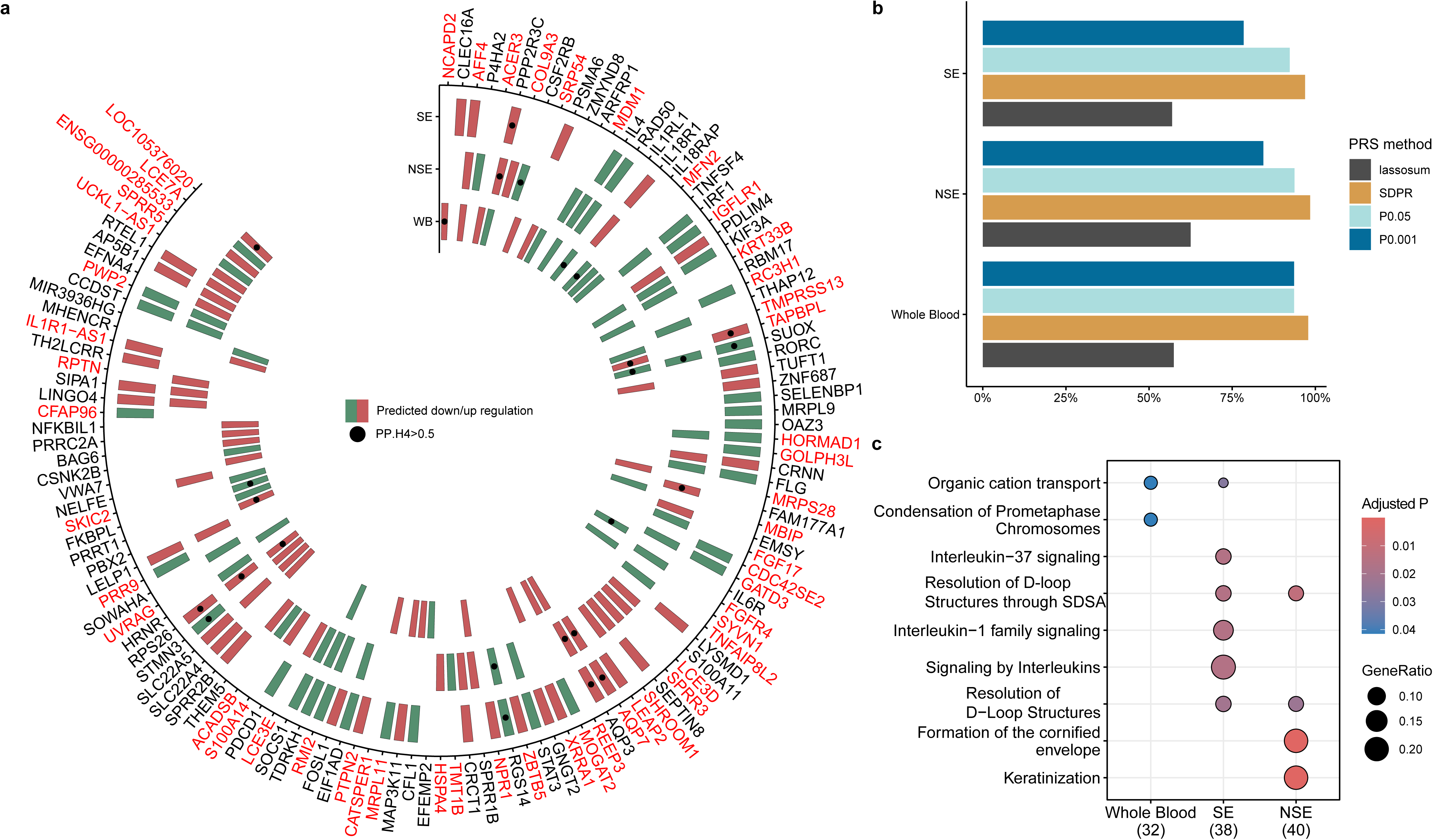
Transcriptome-wide association analysis of atopic dermatitis in 3 different tissues. (a) TWAS results by OTTERS using GWAS and *cis*-eQTL summary statistics for WB, SE and NSE in GTEx V8. Genes colored in red represent TWAS risk genes not identified in TWAS atlas^28^ and the latest published AD TWAS^16^. Each cell represents a gene, with red color reflecting genetically predicted overexpression and green color reflecting genetically predicted underexpression. Each dot indicates significant colocalization defined by a PP.H4>0.5. (b) Percentage of significant genes in each tissue for each PRS method. (c) Pathway enrichment analysis for the significant gene lists per tissue using the Reactome pathways. SE, sun exposed; NSE, not sun exposed; PP.H, posterior probability hypothesis 4; PRS, polygenic risk score.

Given the different assumptions of each PRS method regarding the polygenic architecture, we examined the number of TWAS risk genes each method identified. Lassosum, a methodology reporting large mean squared errors in non-sparse architectures^48^, reported the lowest percentage of identified TWAS risk genes (Fig. 1b). In contrast, the SDPR method, a highly adaptive Dirichlet process prior to model the genetic architecture was declared as the best performing method in all 3 tissues with significant differences from lassosum (Table 1). Notably, P+T methods performed similarly to SDPR apart from the P-value<0.01 method in SE and NSE tissues (Table 1). Further pathway enrichment analyses highlighted tissue-specific functional annotations, referring to significant associations of NSE risk genes in keratinization-related pathways (Fig. 1c). WB risk genes showed an association for cation transporting given the detection of *SLC22A4* and *SLC22A5* genes, while SE risk genes were primarily enriched for inflammatory pathways such as interleukin(IL)-1 family signaling due to the presence of IL18-related signals (Fig. 1c).

**Table 1.**
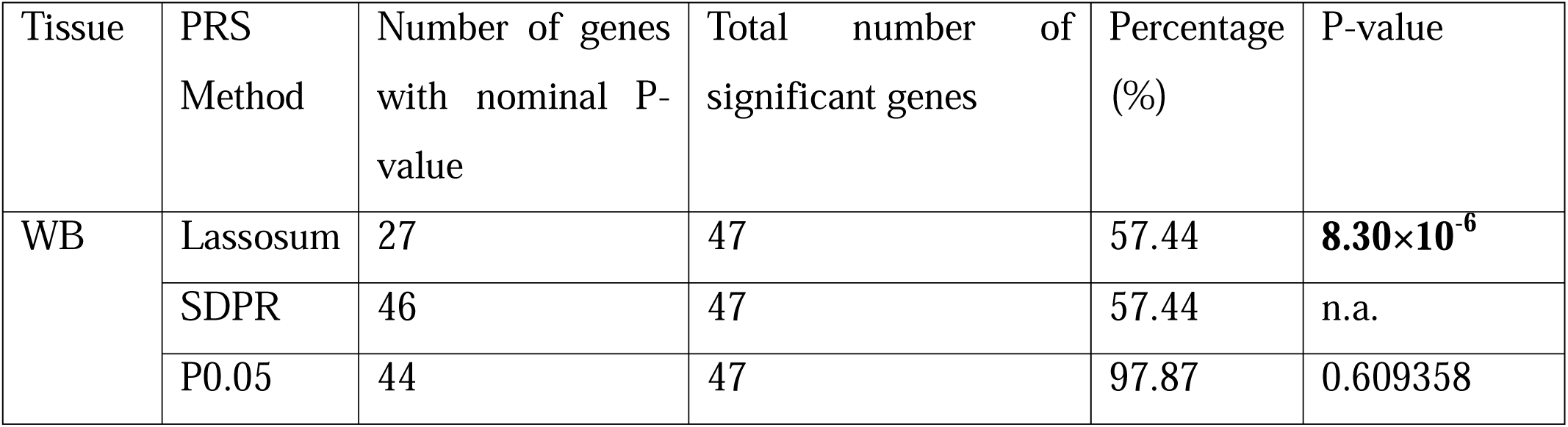
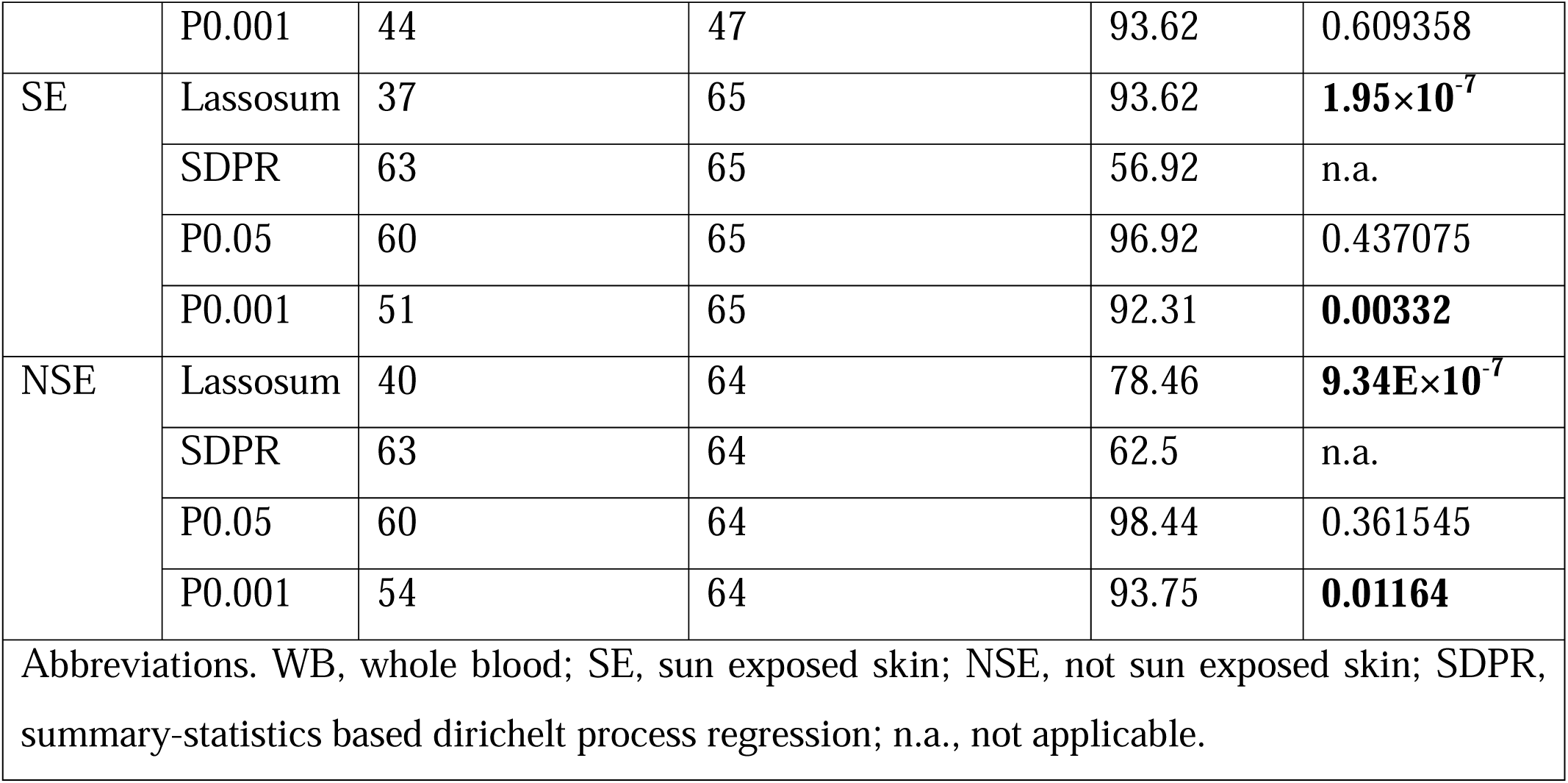
Comparison of polygenic risk score (PRS) method performance in each tissue against SDPR. P-values denoted in bold represent significant associations.

| Tissue | PRS Method | Number of genes with nominal P-value | Total number of significant genes | Percentage (%) | P-value |
| --- | --- | --- | --- | --- | --- |
| WB | Lassosum | 27 | 47 | 57.44 | **8.30×10<sup>-6</sup>** |
|  | SDPR | 46 | 47 | 57.44 | n.a. |
|  | P0.05 | 44 | 47 | 97.87 | 0.609358 |
|  | P0.001 | 44 | 47 | 93.62 | 0.609358 |
| SE | Lassosum | 37 | 65 | 93.62 | **1.95×10<sup>-7</sup>** |
|  | SDPR | 63 | 65 | 56.92 | n.a. |
|  | P0.05 | 60 | 65 | 96.92 | 0.437075 |
|  | P0.001 | 51 | 65 | 92.31 | **0.00332** |
| NSE | Lassosum | 40 | 64 | 78.46 | **9.34E×10<sup>-7</sup>** |
|  | SDPR | 63 | 64 | 62.5 | n.a. |
|  | P0.05 | 60 | 64 | 98.44 | 0.361545 |
|  | P0.001 | 54 | 64 | 93.75 | **0.01164** |
| Abbreviations. WB, whole blood; SE, sun exposed skin; NSE, not sun exposed skin; SDPR, summary-statistics based dirichelt process regression; n.a., not applicable. |  |  |  |  |  |

### Validation of TWAS risk genes

We applied colocalization under a single causal variant assumption to prioritize TWAS risk genes. Colocalization at a PP.H4>0.5 threshold reported significant associations at well-established risk loci, including *RPS26* in all 3 tissues, *RGS14*, *AQP3* and *AQP7* in SE and NSE and *IL18RAP* in WB (Fig. 1a; Table S4). Two novel genes were also prioritized through colocalization, including *TAPBPL* in SE and WB and *NCAPD2* in WB. Tapasin is a vital member of antigen presentation mediating peptide loading in the major histocompatibility complex (MCH) class I molecules, and regulates the activity of T cells. *In vivo* inhibition of *TABPL* ameliorated inflammatory signals in experimental autoimmune encephalomyelitis (EAE) mice, including both the proportions of CD4+ and CD8+ T cells, as well as increased the number of Tregs in the spleen and brain, thus suggesting its potential targeting in T cell-mediated disorders^49^. Conversely, *NCAPD2* has a prominent role in chromosome condensation and has been identified as a risk locus for Alzheimer’s disease^50^. Ablation of *Serpinb3a* in AD mouse models and early exposure to allergens resulted in overexpression of *NCAPD2*, suggesting a possible role of cell cycle pathways in the cutaneous allergic reaction^51^.

### Differential expression meta-analysis

We conducted a meta-analysis of publicly available RNA-seq data to assess the overlap between genetically predicted gene expression transcriptional activity derived from environmental factors. We identified 5 datasets (Table 2) and selected 88 lesional skin biopsies against 98 skin biopsies from healthy controls (Table S5). We observed significant between-study heterogeneity (Fig. S4) and thus applied a random effects meta-analysis in 24545 different transcripts. Among 462 differentially expressed genes (Fig. 2a; Table S6), 8 overlapped with TWAS risk genes. In particular, *RORC* (log_2_FC=-1,2), *TMT1B* (log_2_FC =-2.6) and *MOGAT2* (log_2_FC =-1.62) were repressed in meta-analysis and showed a negative sign in TWASs, while *FOSL1* (log_2_FC =2.12) and *SPRR2B* (log_2_FC =4.93) were upregulated. Discordant signs were observed in *LCE7A*, *CSF2RB* and *SPRR5*. *FOSL1* belongs to the AP1 transcription factor family that orchestrates *FLG* (log_2_FC_meta-analysis_=-0.48) downregulation in *in vitro* models^52^ and associated with erythema skin manifestation^53^. Repression of *TMT1B* methyltransferase expression, a novel TWAS risk gene in NSE (Fig. 1a), has been previously associated with size reduction of malignancies in several cancers^54^.

**Fig. 2.**
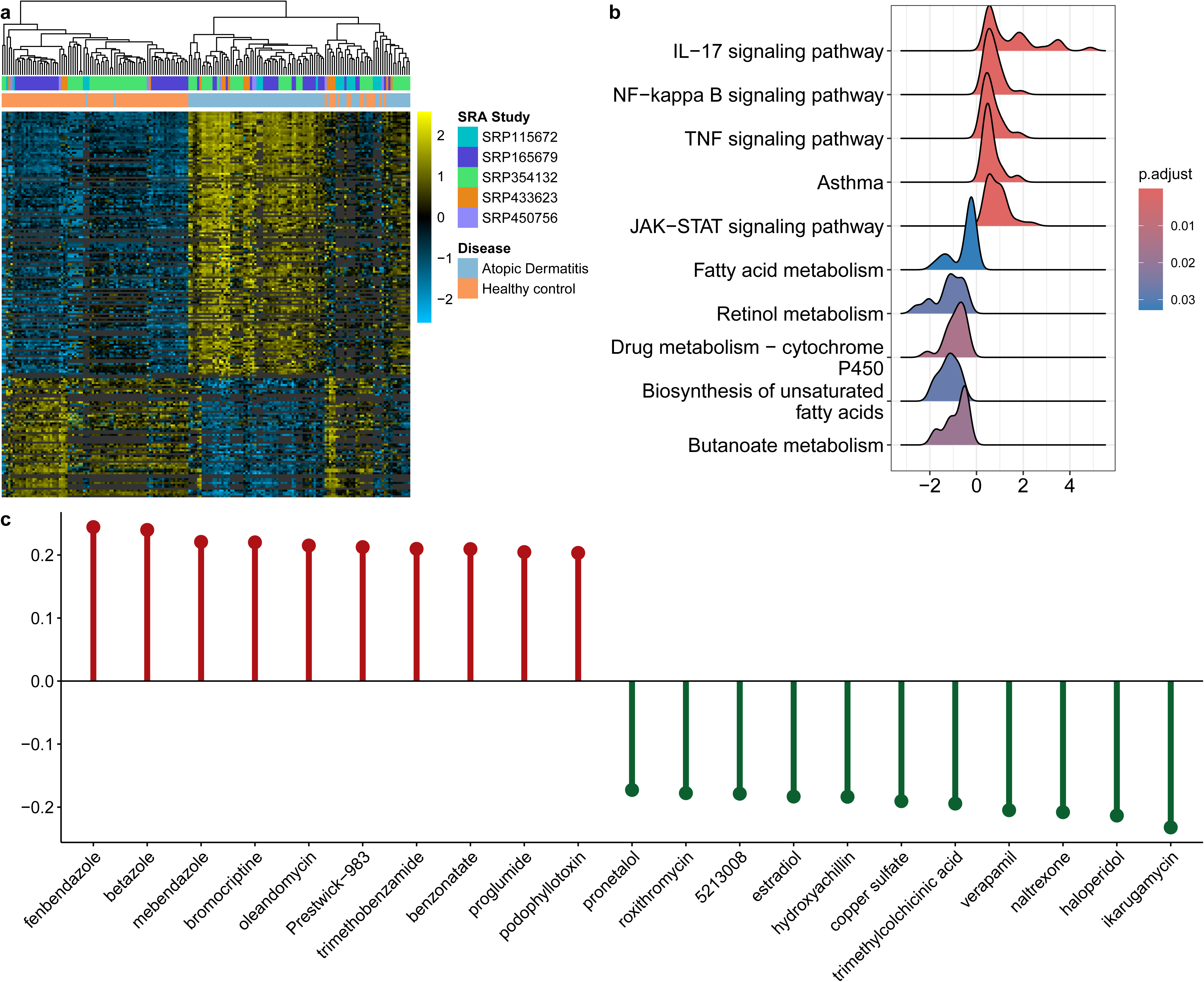
Transcriptomic meta-analysis provides therapeutic opportunities for AD. (a) Heatmap of expression profiles for all differentially expressed genes (n=462) in the meta-analysis. (b) Ridgeplot of selected pathways derived from the GSEA in the KEGG pathways database. The x-axis refers to the normalized enrichment score. (c) Cosine similarity between the meta-analyzed AD signatures and drug signatures retrieved from the combination of the Connectivity map build 02 and LINCS L1000 datasets. Red color indicates positive cosine similarity and green color represents negative cosine similarity.

**Table 2.** Summary of datasets used in the current study.

| Study | Accession ID | Dataset | Sequencing technology | Strand specificity | Average spot length | Cases | Controls |
| --- | --- | --- | --- | --- | --- | --- | --- |
| Oetjen LK et al., 2017<sup>72</sup> | GSE237920 | SRP450756 | Single | Unstranded | 50 | 4 | 4 |
| Kim M et al., 2024<sup>73</sup> | GSE230200 | SRP433623 | Single | Reverse stranded | 101 | 10 | 8 |
| Blunder et al., 2018<sup>74</sup> | GSE102628 | SRP115672 | Single | Unstranded | 64 | 12 | 9 |
| Tsoi LC et al., 2019<sup>75</sup> | GSE121212 | SRP165679 | Paired | Reverse stranded | 252 | 27 | 38 |
| Hu et al., 2023<sup>76</sup> | GSE193309 | SRP354132 | Paired | Unstranded | 302 | 35 | 39 |

The meta-analysis results further provided an AD-like transcriptome at the pathway level, including the overexpression of asthma-related genes and inflammatory pathways of TNF and NF-kB signaling, while metabolic pathways were repressed (Fig. 2b; Table S7). We next attempted to identify drug signatures that have similar and/or opposing effects with the meta-analysis summary statistics through the intersection of CMAP build 02 and LINCS L1000 databases. Drugs with positive cosine similarity suggest similar transcriptomic profile to AD, while drugs reporting a negative cosine similarity show an opposite expression direction.

Fenbendazole and betazole belonged in the top enriched drugs with positive cosine similarity, while ikarugamycin and haloperidol were the most enriched drugs for negative cosine similarity (Fig. 2c). On the one hand, ikarugamycin has displayed potent anti-inflammatory effects by inducing the activation of A20 and thus limiting NF-kB activity^55^. On the contrary, haloperidol is a well-known antipsychotic drug^56^, complementing previous work establishing a strong link between AD and neuropsychiatric traits^57^.

### Co-expression network analysis

We finally sought to map the regulatory landscape and potential co-expression interactions of TWAS risk signals in the transcriptomic signature of cutaneous AD. We required only paired-end RNA-seq datasets for WGCNA and identified 43 outliers using the ROBPCA algorithm^42^ (Fig. S5). A total of 24546 transcripts (Fig. S6) from 98 samples (n_cases_=39, n_controls_=59) were submitted in the WGCNA, with a soft-power thresholding set at 16 (Fig. S7). We identified 48 co-expression modules excluding the gray module (Fig. S8; Table S8), with 24 being significantly associated with AD risk (14 positive, 10 negative; Fig. 3a; Table S9). The largest module (ME1) contained 4085 genes, while ME48 reported the lowest number of genes (n=26; Table S8). All modules met quality criteria (Fig. 3b) and were prioritized for identification of hub genes (MM>0.8 and GS>0.2). In total, 16 unique TWAS risk genes were also identified as hub genes in the respective co-expression modules, with *FOSL1* as an exemplar (Fig. 3c). Other examples include *CFL1*, *AQP3,* the novel TWAS risk gene *AQP7*, small proline rich proteins 1B and 2B, and *RORC* (Fig. 3c). We submitted all 16 non-overlapping genes in the DGIdb platform to characterize potential novel drug-gene interactions (Table S10). The highest interaction score was found for *MAP3K11* and CEP-1347, a semi-synthetic compound with neuroprotective activity in rodents^58^. Sargramostim interacts with *CSF2RB* to induce JAK2/STAT1-STAT3 signaling pathway and stimulate T cell differentiation. Previous studies have suggested that over-expression of *CSF2R*, the target of sargramostim, may increase wound healing in keratinocytes and therefore clearance of inflammation in keratinocytes^59^. Additional drug-gene interactions refer to *CSF2RB* and mavrilimumab, a novel agent for rheumatoid arthritis^60^, and *CFL1* with clofibrate, a lipid-lowering agent previously administered in AD patients^61^.

**Fig. 3.**
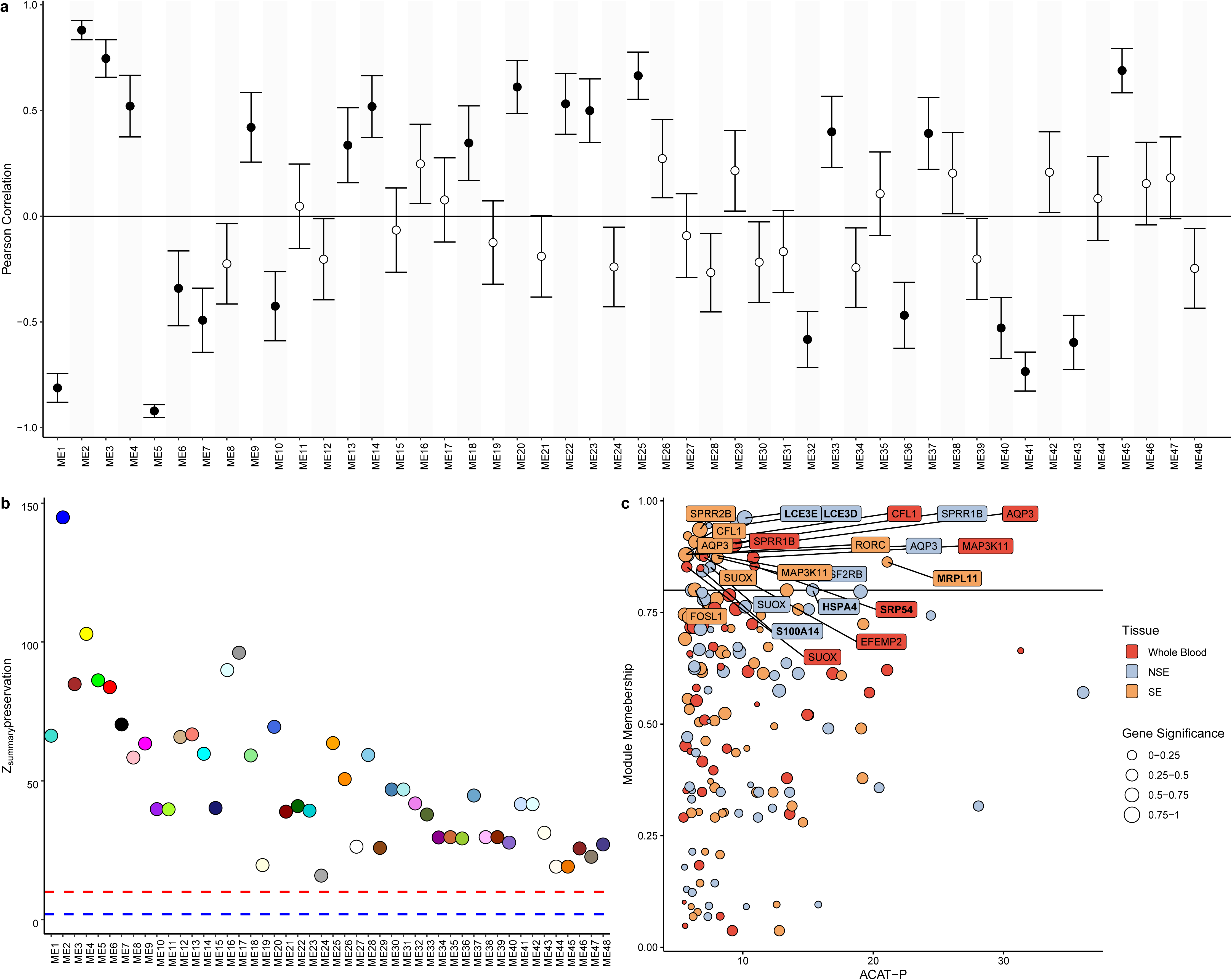
Triangulating co-expression evidence to prioritize TWAS risk genes. (a) Pearson correlation coefficients from all identified modules. Filled dots represent significant correlations with AD. (b) Module preservation Z-summary statistics across 48 co-expression modules. The blue dashed line shows Z-summary≤2 (low preservation), while the red dashed line shows 2≤Z-summary≤10 (moderate preservation). Modules with Z-summary>10 are considered as highly preserved. (c) Comparison of module membership and gene significance with TWAS risk genes. Size of each dot is scaled according to the gene significance. Genes with module membership>0.8 and gene significance>0.2 are annotated. Genes in bold represent TWAS risk genes not identified in TWAS atlas^28^ and the latest published AD TWAS^16^.

## Discussion

Here, we conducted a large-scale TWAS of AD focusing on 3 tissues representative of systemic and localized AD pathophysiology. Despite previous research has identified a larger number of enriched tissue expressions in AD, we focused on WB, SE and NSE to represent the systemic and localized AD pathophysiology respectively. We detected 126 unique TWAS risk signals across all tissues, unveiling 53 novel gene associations compared to previous TWASs in AD. Assuming a single shared causal variant between AD and *cis*-eQTL summary statistics, we validated two novel risk genes for AD (*TAPBL* and *NCAPD2*), apart from validation of well-known risk genes for AD (e.g., *IL6R*, *IL18R1*). The identification of *TAPBPL* as a TWAS risk gene highlights its role in T-cell modulation. This finding aligns with prior studies showing its involvement in antigen presentation, suggesting a novel therapeutic target for immune-related aspects of AD. By integrating TWAS and co-expression networks, we identified 16 key hub genes, such as *FOSL1*, that regulate skin inflammation and keratinization—pathways central to AD pathogenesis. These findings not only expand our understanding of AD pathophysiology but also provide actionable targets for drug development, such as *AQP7* for skin barrier repair and *CSF2RB* for inflammation modulation.

Compared to previous TWAS conducted in AD, we employed a larger GWAS comprised of more than 800000 participants of European ancestry, as well as a novel method for predicting genetically regulated gene expression. In particular, OTTERS integrates multiple genetic architecture assumptions for expression variations through PRS models to estimate gene expression regulation. Given the increased computational costs in applying PRS-CS, we prioritized P+T methods, SDPR and lassosum given their complementary performance in relevant simulations^21^. Consistent to the original OTTERS implementation in cardiovascular disease^21^, we observed a superior performance of SDPR across all PRS models in each tissue (Table 1). The highest P+T approach (P-value<0.05) further detected a comparable number of TWAS risk signals across tissues (Table 1). However, more stringent P-value thresholds (P-value<0.001), excluding a larger number of *cis*-eQTLs at Stage I, yielded significant differences compared to the superior SDPR in SE and NSE tissues (Table 1). On the contrary, lassosum, a PRS model based on penalized regression with improved performance over sparse architectures^62^, demonstrated poor effectiveness in retrieving TWAS risk signals (Fig. 1b). These findings support the notion that AD risk is less likely driven by sparse risk loci (e.g., protein-coding variants in genes relevant to skin barrier integrity), rather than a combination of functionally relevant loci in a wide variety of AD-related biological pathways.

Indeed, previous gene prioritization efforts in AD remained inconclusive at well-established risk loci, where the *FLG* protein-coding gene was ranked sixth in the 1q21.3 locus^13^. Our TWAS findings corroborate previously reported associations including IL18-related genes such as *IL18RAP* and *IL18R1* and *HRNR* (Fig. 1a). Additionally, novel TWAS risk genes encompass a broad spectrum of functional understanding, incorporating genes with limited studies in allergic, immune and skin functions (e.g., *PWP2*, *ACADSB*, *ZMYND8*) as well as genes with established roles in AD-relevant pathways (e.g., *TMPRSS13* in skin barrier integrity^63^, *TAPBPL* in antigen presentation^49^ and *XRRA1* as a risk locus for asthma in African American participants^64^ among others). Integration of co-expression network and TWAS risk signals further enabled us to highlight hub genes that may modulate disease risk in skin (Fig. 3c). For instance, *CFL1* has been previously associated with AD through TWAS, but no functional insights have been gained for its role in AD pathogenesis. Cofilin 1 is an intracellular actin-modulating protein that has been validated as a potential biomarker for AD^65^ and associated with *FLG* deficiency through proteomic assays^66^. On the other hand, common *CSF2RB* variants contribute to the AD risk^67^, while ablation of *Csf2rb* inhibited extensive allergic reactions and accumulation of T_H_2 cells^68^. Future experimental validation is required for several of these genes, as well as their potential assessment as candidate drugs.

Our study has caveats. We limited our analyses to participants of European ancestry thereby restricting the generalizability of our results. For example, Asian patients with AD demonstrate a significantly higher proportion of T_H_17-related signatures than European patients, hence suggesting an ancestry-specific genetic component^69,70^. Moreover, we focused on *cis*-eQTL variants mapped ±1MB around the transcription start and end sites, excluding *trans*-eQTLs and variants influencing gene expression via long-range chromatin interaction. Additionally, we restricted our analysis to three tissues, namely WB, SE and NSE, as they represent key systemic and localized pathophysiological sites of AD. Hence, some gene-disease associations in a tissue-specific matter may have been overlooked, as is the case with *OVOL1* expression, a known risk locus in AD, reporting significant results only in Epstein-Barr virus transformed lymphocytes^14,16^. Future work could incorporate a larger number of tissues, as well as cell-specific cis-eQTLs that regulate inflammatory processes in AD. In addition, although TWASs mitigate concerns of reverse causality, they cannot distinguish pleiotropic associations between *cis*-eQTLs and GWASs and genuinely causal genes. Lastly, our meta-analysis approach did not incorporate clinical covariates due to inconsistent reporting across the included datasets, which may influence expression profiles. To address this as well as significant heterogeneity observed (Fig. S4), we conducted a random-effects meta-analysis, yielding robust findings both at the gene level (Fig. 2a) and the pathway level (Fig. 2b).

Overall, we leverage *cis*-eQTLs from 3 representative tissues to identify novel risk genes in AD through TWASs. We report 53 novel TWAS risk genes with previously unidentified roles in AD pathogenesis. Through the integration of TWASs with RNA-seq, we highlight 16 genes (Fig. 3c) that could be prioritized for follow-up studies and functional experiments.

## Data Availability

All data produced in the present study are presented in the supplementary files.

## Acknowledgements

CA was financially funded by the “Andreas Mentzelopoulos Foundation”.

## Author Contributions

Conceptualization: CA; Methodology: CA, DM, PX; Investigation: CA, DM, AP, AA, PX; Visualization: CA, DM; Supervision: YV; Writing-Original Draft: CA, DM; Writing-review and editing: CA, DM, AP, AA, PX, GKG, SG, AT, StG, KG, YV

## Data and materials availability

Summary statistics for atopic dermatitis can be downloaded from the GWAS catalog (https://www.ebi.ac.uk/gwas/). Cis-eQTL summary statistics can be downloadad from the GTEx V8 portal (https://gtexportal.org/home/) and the eQTL catalogue (https://www.ebi.ac.uk/eqtl/).

## Supplementary Figures

Fig S1. QQ plot of ACAT P-values pre (left) and post (right) adjustment for genomic control in GTEx V8 Whole Blood.

Fig S2. QQ plot of ACAT P-values pre (left) and post (right) adjustment for genomic control in GTEx V8 Sun exposed skin.

Fig S3. QQ plot of ACAT P-values pre (left) and post (right) adjustment for genomic control in GTEx V8 Not Sun exposed skin.

Fig S4. QQ plot of the Cochran’s test for the transcriptomic meta-analysis.

Fig S5. Principal component analysis (PCA) pre (left) and post (right) outlier correction based on the ROBPCA algorithm.

Fig S6. Heatmap of transcripts clustered during the WGCNA analysis. Transcripts falling in the gray module (ME0) were excluded from the heatmap.

Fig S7. Determination of the soft-thresholding power for WGCNA. Fig. S8. Cluster dendrogram and colored co-expression modules.

## Supplementary Tables

Table S1. Transcriptome-wide association study results in GTEx V8 Whole Blood. Table S2. Transcriptome-wide association study results in GTEx V8 Sun Exposed skin.

Table S3. Transcriptome-wide association study results in GTEx V8 Not Sun Exposed skin. Table S4. Colocalization results for TWAS risk genes in each tissue.

Table S5. Sample IDs included in the meta-analysis Table S6. Transcriptomic meta-analysis results.

Table S7. Gene-set enrichment analysis of transcriptomic meta-analysis with KEGG pathways. Table S8. Co-expression modules in paired-end RNA-seq datasets.

Table S9. Pearson correlation coefficient between module expressions and atopic dermatitis. Table S10. Drug-gene interactions for prioritized TWAS risk genes based on the DGIdb platform.

## References

1. Hill DA, Spergel JM. The atopic march: Critical evidence and clinical relevance. Ann Allergy Asthma Immunol 2018; 120:131–7.

2. Eyerich K, Novak N. Immunology of atopic eczema: overcoming the Th1/Th2 paradigm. Allergy 2013; 68:974–82.

3. Hill DA, Spergel JM. The atopic march: Critical evidence and clinical relevance. Ann Allergy Asthma Immunol 2018; 120:131–7.

4. Budu-Aggrey A, Kilanowski A, Sobczyk MK, et al. European and multi-ancestry genome-wide association meta-analysis of atopic dermatitis highlights importance of systemic immune regulation. Nat Commun 2023; 14:6172.

5. Antonatos C, Grafanaki K, Georgiou S, et al. Disentangling the complexity of psoriasis in the post-genome-wide association era. Genes Immun 2023; 24:236–47.

6. Nica AC, Dermitzakis ET. Expression quantitative trait loci: present and future. Phil Trans R Soc B 2013; 368:20120362.

7. Gamazon ER, Wheeler HE, Shah KP, et al. A gene-based association method for mapping traits using reference transcriptome data. Nat Genet 2015; 47:1091–8.

8. Mai J, Lu M, Gao Q, et al. Transcriptome-wide association studies: recent advances in methods, applications and available databases. Commun Biol 2023; 6:899.

9. Zhu Z, Zhang F, Hu H, et al. Integration of summary data from GWAS and eQTL studies predicts complex trait gene targets. Nat Genet 2016; 48:481–7.

10. Gusev A, Ko A, Shi H, et al. Integrative approaches for large-scale transcriptome-wide association studies. Nat Genet 2016; 48:245–52.

11. Zhou D, Jiang Y, Zhong X, et al. A unified framework for joint-tissue transcriptome-wide association and Mendelian randomization analysis. Nat Genet 2020; 52:1239–46.

12. GTEx Consortium. Genetic effects on gene expression across human tissues. Nature 2017; 550:204–13.

13. Sobczyk MK, Richardson TG, Zuber V, et al. Triangulating Molecular Evidence to Prioritize Candidate Causal Genes at Established Atopic Dermatitis Loci. Journal of Investigative Dermatology 2021; 141:2620–9.

14. Song J, Kim D, Lee S, et al. Integrative transcriptome-wide analysis of atopic dermatitis for drug repositioning. Commun Biol 2022; 5:615.

15. The GTEx Consortium, Aguet F, Anand S, et al. The GTEx Consortium atlas of genetic regulatory effects across human tissues. Science 2020; 369:1318–30.

16. Wu H, Ke X, Huang W, et al. Multitissue Integrative Analysis Identifies Susceptibility Genes for Atopic Dermatitis. Journal of Investigative Dermatology 2023; 143:602–611.e14.

17. Budu-Aggrey A, Kilanowski A, Sobczyk MK, et al. European and multi-ancestry genome-wide association meta-analysis of atopic dermatitis highlights importance of systemic immune regulation. Nat Commun 2023; 14:6172.

18. Song J, Kim D, Lee S, et al. Integrative transcriptome-wide analysis of atopic dermatitis for drug repositioning. Commun Biol 2022; 5:615.

19. Byrska-Bishop M, Evani US, Zhao X, et al. High-coverage whole-genome sequencing of the expanded 1000 Genomes Project cohort including 602 trios. Cell 2022; 185:3426–3440.e19.

20. Xu Y, He C, Fan J, et al. A multi-modal framework improves prediction of tissue-specific gene expression from a surrogate tissue. eBioMedicine 2024; 107:105305.

21. Dai Q, Zhou G, Zhao H, et al. OTTERS: a powerful TWAS framework leveraging summary-level reference data. Nat Commun 2023; 14:1271.

22. Privé F, Vilhjálmsson BJ, Aschard H, Blum MGB. Making the Most of Clumping and Thresholding for Polygenic Scores. The American Journal of Human Genetics 2019; 105:1213–21.

23. Mak TSH, Porsch RM, Choi SW, et al. Polygenic scores via penalized regression on summary statistics. Genetic Epidemiology 2017; 41:469–80.

24. Ge T, Chen C-Y, Ni Y, et al. Polygenic prediction via Bayesian regression and continuous shrinkage priors. Nat Commun 2019; 10:1776.

25. Zhou G, Zhao H. A fast and robust Bayesian nonparametric method for prediction of complex traits using summary statistics. PLoS Genet 2021; 17:e1009697.

26. Liu Y, Chen S, Li Z, et al. ACAT: A Fast and Powerful p Value Combination Method for Rare-Variant Analysis in Sequencing Studies. The American Journal of Human Genetics 2019; 104:410–21

27. Devlin B, Roeder K, Wasserman L. Genomic Control, a New Approach to Genetic-Based Association Studies. Theoretical Population Biology 2001; 60:155–66.

28. Lu M, Zhang Y, Yang F, et al. TWAS Atlas: a curated knowledgebase of transcriptome-wide association studies. Nucleic Acids Res 2023; 51:D1179–87.

29. Xiropotamos P, Papageorgiou F, Manousaki H, et al. aPEAch: Automated Pipeline for End-to-End Analysis of Epigenomic and Transcriptomic Data. Biology 2024; 13:492.

30. Dobin A, Davis CA, Schlesinger F, et al. STAR: ultrafast universal RNA-seq aligner. Bioinformatics 2013; 29:15–21.

31. Li H, Handsaker B, Wysoker A, et al. The Sequence Alignment/Map format and SAMtools. Bioinformatics 2009; 25:2078–9.

32. Liao Y, Smyth GK, Shi W. featureCounts: an efficient general purpose program for assigning sequence reads to genomic features. Bioinformatics 2014; 30:923–30.

33. Wang L, Wang S, Li W. RSeQC: quality control of RNA-seq experiments. Bioinformatics 2012; 28:2184–5.

34. Ritchie ME, Phipson B, Wu D, et al. limma powers differential expression analyses for RNA-sequencing and microarray studies. Nucleic Acids Res 2015; 43:e47.

35. Villatoro-García JA, Martorell-Marugán J, Toro-Domínguez D, et al. DExMA: An R Package for Performing Gene Expression Meta-Analysis with Missing Genes. Mathematics 2022; 10:3376.

36. Mancuso CA, Canfield JL, Singla D, Krishnan A. A flexible, interpretable, and accurate approach for imputing the expression of unmeasured genes. Nucleic Acids Research 2020; 48:e125–e125.

37. Subramanian A, Narayan R, Corsello SM, et al. A Next Generation Connectivity Map: L1000 Platform and the First 1,000,000 Profiles. Cell 2017; 171:1437–1452.e17.

38. Pickering A. ccmap: Combination Connectivity Mapping. 2024. R package version 1.32.0.

39. Langfelder P, Horvath S. WGCNA: an R package for weighted correlation network analysis. BMC Bioinformatics 2008; 9:559.

40. Antonatos C, Georgakilas GK, Evangelou E, Vasilopoulos Y. Transcriptomic meta-analysis characterizes molecular commonalities between psoriasis and obesity. Genes Immun 2024; 25:179–87.

41. Love MI, Huber W, Anders S. Moderated estimation of fold change and dispersion for RNA-seq data with DESeq2. Genome Biol 2014; 15:550.

42. Hubert M, Rousseeuw PJ, Vanden Branden K. ROBPCA: A New Approach to Robust Principal Component Analysis. Technometrics 2005; 47:64–79.

43. Cannon M, Stevenson J, Stahl K, et al. DGIdb 5.0: rebuilding the drug-gene interaction database for precision medicine and drug discovery platforms. Nucleic Acids Res 2024; 52:D1227–35.

44. Hara Chikuma M, Sugiyama Y, Kabashima K, et al. Involvement of aquaporin 7 in the cutaneous primary immune response through modulation of antigen uptake and migration in dendritic cells. FASEB j 2012; 26:211–8.

45. McLeod O, Silveira A, Valdes-Marquez E, et al. Genetic loci on chromosome 5 are associated with circulating levels of interleukin-5 and eosinophil count in a European population with high risk for cardiovascular disease. Cytokine 2016; 81:1–9.

46. Pividori M, Schoettler N, Nicolae DL, et al. Shared and distinct genetic risk factors for childhood-onset and adult-onset asthma: genome-wide and transcriptome-wide studies. The Lancet Respiratory Medicine 2019; 7:509–22.

47. Bhattacharya N, Sato WJ, Kelly A, et al. Epidermal Lipids: Key Mediators of Atopic Dermatitis Pathogenesis. Trends Mol Med 2019; 25:551–62.

48. Yang S, Zhou X. Accurate and Scalable Construction of Polygenic Scores in Large Biobank Data Sets. Am J Hum Genet 2020; 106:679–93.

49. Lin Y, Cui C, Su M, et al. Identification of TAPBPL as a novel negative regulator of T-cell function. EMBO Mol Med 2021; 13:e13404.

50. Pang D, Yu S, Yang X. A mini-review of the role of condensin in human nervous system diseases. Front Mol Neurosci 2022; 15:889796.

51. Sivaprasad U, Kinker KG, Ericksen MB, et al. SERPINB3/B4 Contributes to Early Inflammation and Barrier Dysfunction in an Experimental Murine Model of Atopic Dermatitis. Journal of Investigative Dermatology 2015; 135:160–9.

52. Ahn SS, Yeo H, Jung E, et al. FRA1:c-JUN:HDAC1 complex down-regulates filaggrin expression upon TNFα and IFNγ stimulation in keratinocytes. Proc Natl Acad Sci U S A 2022; 119:e2123451119.

53. Sekita A, Kawasaki H, Fukushima-Nomura A, et al. Multifaceted analysis of cross-tissue transcriptomes reveals phenotype–endotype associations in atopic dermatitis. Nat Commun 2023; 14:6133.

54. Denford SE, Wilhelm BT. Defining the elusive oncogenic role of the methyltransferase TMT1B. Front Oncol 2023; 13:1211540.

55. Malcomson B, Wilson H, Veglia E, et al. Connectivity mapping (ssCMap) to predict A20-inducing drugs and their antiinflammatory action in cystic fibrosis. Proc Natl Acad Sci U S A 2016; 113:E3725–3734.

56. Dold M, Samara MT, Li C, et al. Haloperidol versus first-generation antipsychotics for the treatment of schizophrenia and other psychotic disorders. Cochrane Database Syst Rev 2015; 1:CD009831.

57. Antonatos C, Pontikas A, Akritidis A, et al. A genome-wide pleiotropy study between atopic dermatitis and neuropsychiatric disorders. 2024. doi:10.1101/2024.10.30.24316209.

58. Parkinson Study Group PRECEPT Investigators. Mixed lineage kinase inhibitor CEP-1347 fails to delay disability in early Parkinson disease. Neurology 2007; 69:1480–90.

59. Mascia F, Cataisson C, Lee T-C, et al. EGFR regulates the expression of keratinocyte-derived granulocyte/macrophage colony-stimulating factor in vitro and in vivo. J Invest Dermatol 2010; 130:682–93.

60. Burmester GR, Feist E, Sleeman MA, et al. Mavrilimumab, a human monoclonal antibody targeting GM-CSF receptor-α, in subjects with rheumatoid arthritis: a randomised, double-blind, placebo-controlled, phase I, first-in-human study. Ann Rheum Dis 2011; 70:1542–9.

61. Fukaya M, Kimata H. Topical clofibrate improves symptoms in patients with atopic dermatitis and reduces serum TARC levels: a randomized, double-blind, placebo-controlled pilot study. J Drugs Dermatol 2014; 13:259–63.

62. Wheeler HE, Shah KP, Brenner J, et al. Survey of the Heritability and Sparse Architecture of Gene Expression Traits across Human Tissues. PLoS Genet 2016; 12:e1006423.

63. Madsen DH, Szabo R, Molinolo AA, Bugge TH. TMPRSS13 deficiency impairs stratum corneum formation and epidermal barrier acquisition. Biochem J 2014; 461:487–95.

64. Chang X, March M, Mentch F, et al. Genetic architecture of asthma in African American patients. Journal of Allergy and Clinical Immunology 2023; 151:1132–6.

65. Zhou X, Xiao B, Zeng J, et al. Identification of Cofilin-1 as a novel biomarker of atopic dermatitis using iTRAQ quantitative proteomics. J Clin Lab Anal 2022; 36:e24751.

66. Elias MS, Long HA, Newman CF, et al. Proteomic analysis of filaggrin deficiency identifies molecular signatures characteristic of atopic eczema. J Allergy Clin Immunol 2017; 140:1299–309.

67. Grosche S, Marenholz I, Esparza-Gordillo J, et al. Rare variant analysis in eczema identifies exonic variants in DUSP1, NOTCH4 and SLC9A4. Nat Commun 2021; 12:6618.

68. Asquith KL, Ramshaw HS, Hansbro PM, et al. The IL-3/IL-5/GM-CSF Common β Receptor Plays a Pivotal Role in the Regulation of Th2 Immunity and Allergic Airway Inflammation. The Journal of Immunology 2008; 180:1199–206.

69. Chan TC, Sanyal RD, Pavel AB, et al. Atopic dermatitis in Chinese patients shows TH2/TH17 skewing with psoriasiform features. Journal of Allergy and Clinical Immunology 2018; 142:1013–7.

70. Noda S, Suárez-Fariñas M, Ungar B, et al. The Asian atopic dermatitis phenotype combines features of atopic dermatitis and psoriasis with increased TH17 polarization. Journal of Allergy and Clinical Immunology 2015; 136:1254–64.

71. Yazar S, Alquicira-Hernandez J, Wing K, et al. Single-cell eQTL mapping identifies cell type–specific genetic control of autoimmune disease. Science 2022; 376:eabf3041.

72. Oetjen LK, Mack MR, Feng J, et al. Sensory Neurons Co-opt Classical Immune Signaling Pathways to Mediate Chronic Itch. Cell 2017; 171:217–228.e13.

73. Kim M, Renert-Yuval Y, Stepensky P, et al. Sclerotic-Type Cutaneous Chronic Graft-Versus-Host Disease Exhibits Activation of T Helper 1 and OX40 Cytokines. J Invest Dermatol 2024; 144:563–572.e9.

74. Blunder S, Kõks S, Kõks G, et al. Enhanced Expression of Genes Related to Xenobiotic Metabolism in the Skin of Patients with Atopic Dermatitis but Not with Ichthyosis Vulgaris. J Invest Dermatol 2018; 138:98–108.

75. Tsoi LC, Rodriguez E, Degenhardt F, et al. Atopic Dermatitis Is an IL-13-Dominant Disease with Greater Molecular Heterogeneity Compared to Psoriasis. J Invest Dermatol 2019; 139:1480–9.

76. Hu T, Todberg T, Ewald DA, et al. Assessment of Spatial and Temporal Variation in the Skin Transcriptome of Atopic Dermatitis by Use of 1.5 mm Minipunch Biopsies. J Invest Dermatol 2023; 143:612–620.e6.

