## Supplementary material for "Transcriptome-wide analyses delineate the genetic architecture of expression variation in atopic dermatitis": Fig. S

Fig S1. QQ plot of ACAT P-values pre (left) and post (right) adjustment for genomic control in GTEx V8 Whole Blood.

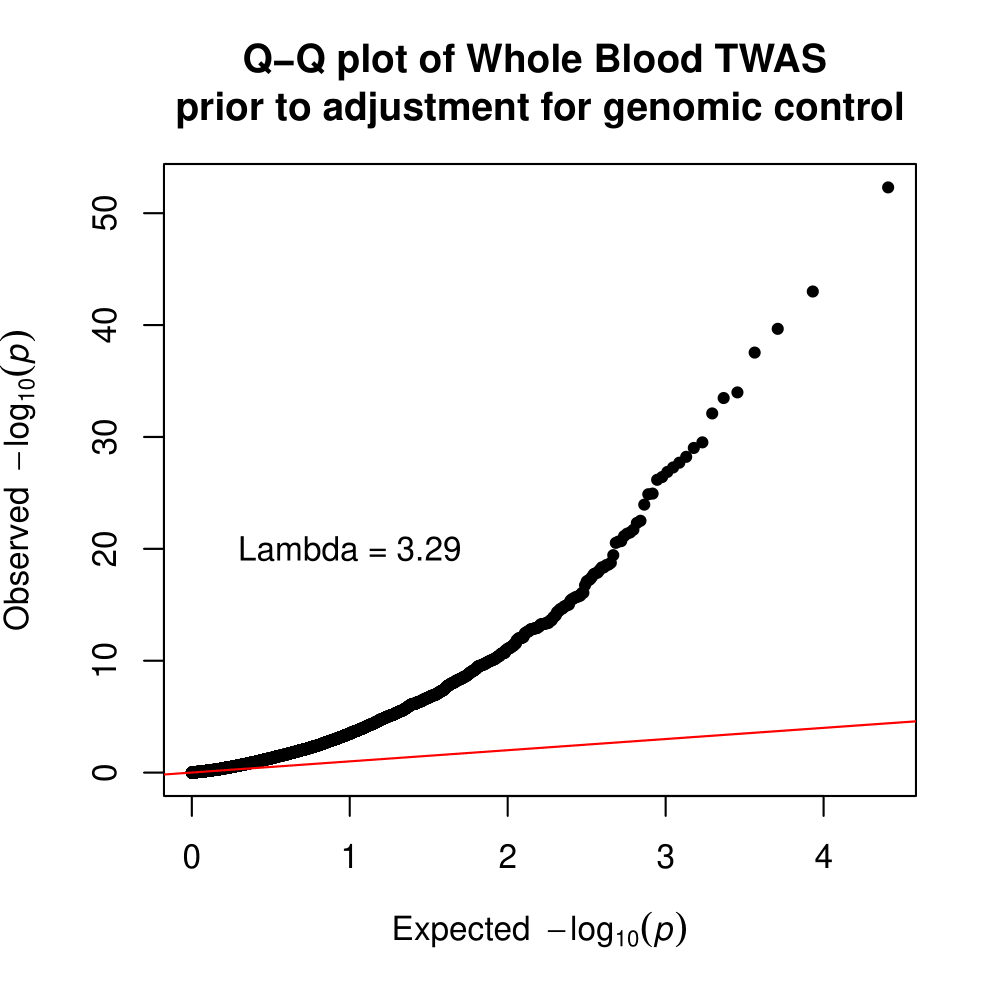

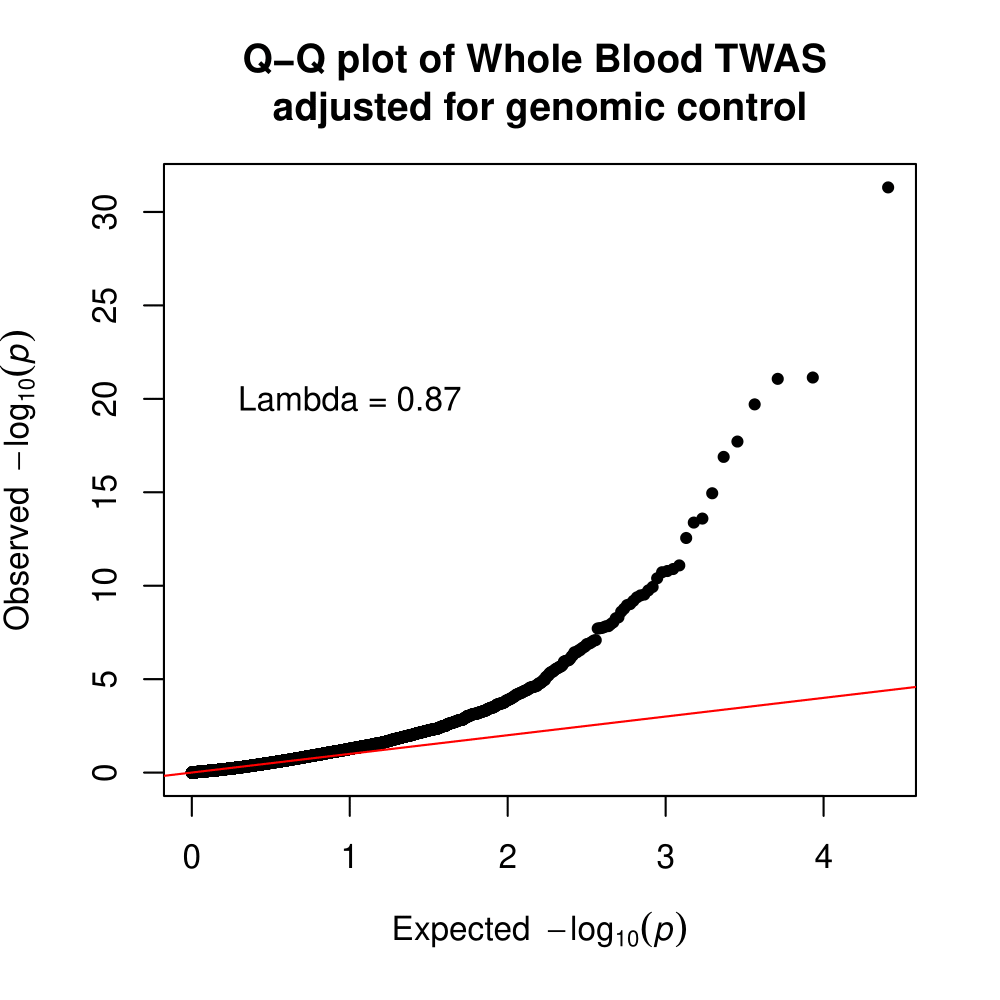

Fig S2. QQ plot of ACAT P-values pre (left) and post (right) adjustment for genomic control in GTEx V8 Sun exposed skin.

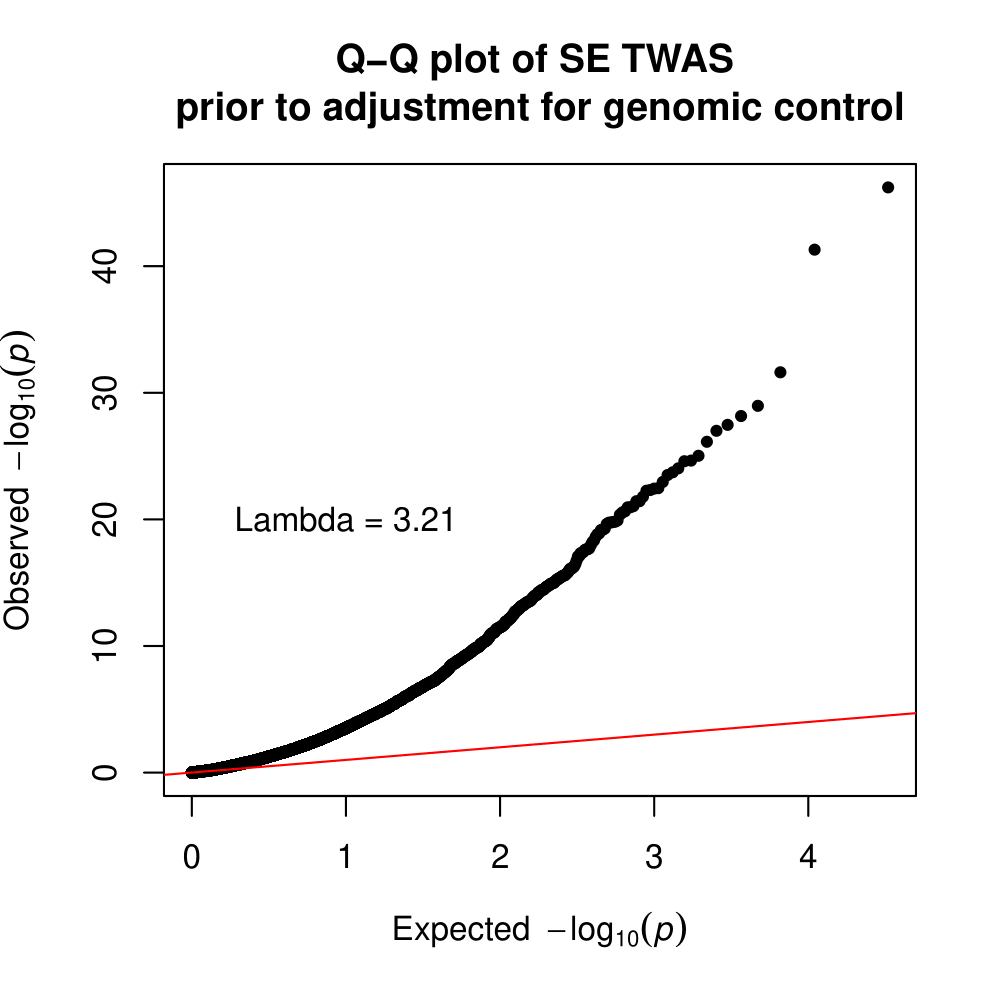

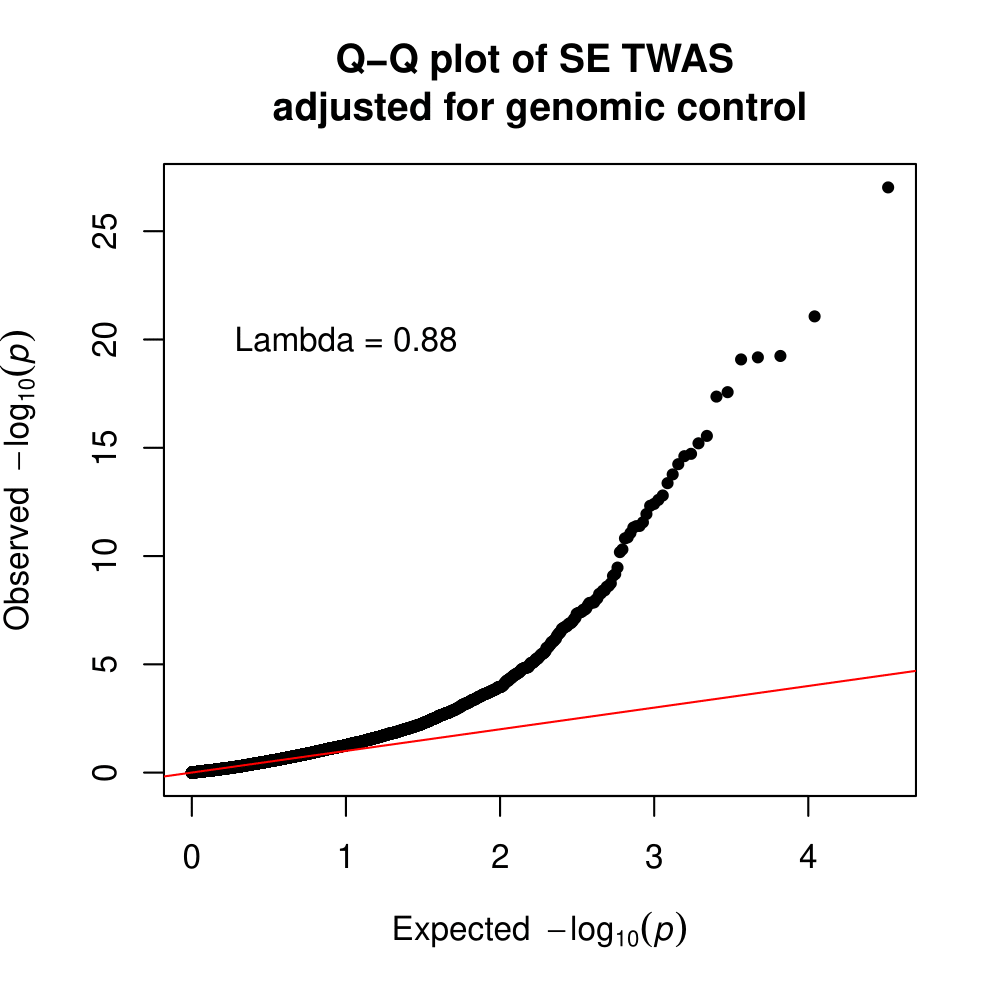

Fig S3. QQ plot of ACAT P-values pre (left) and post (right) adjustment for genomic control in GTEx V8 Not Sun exposed skin.

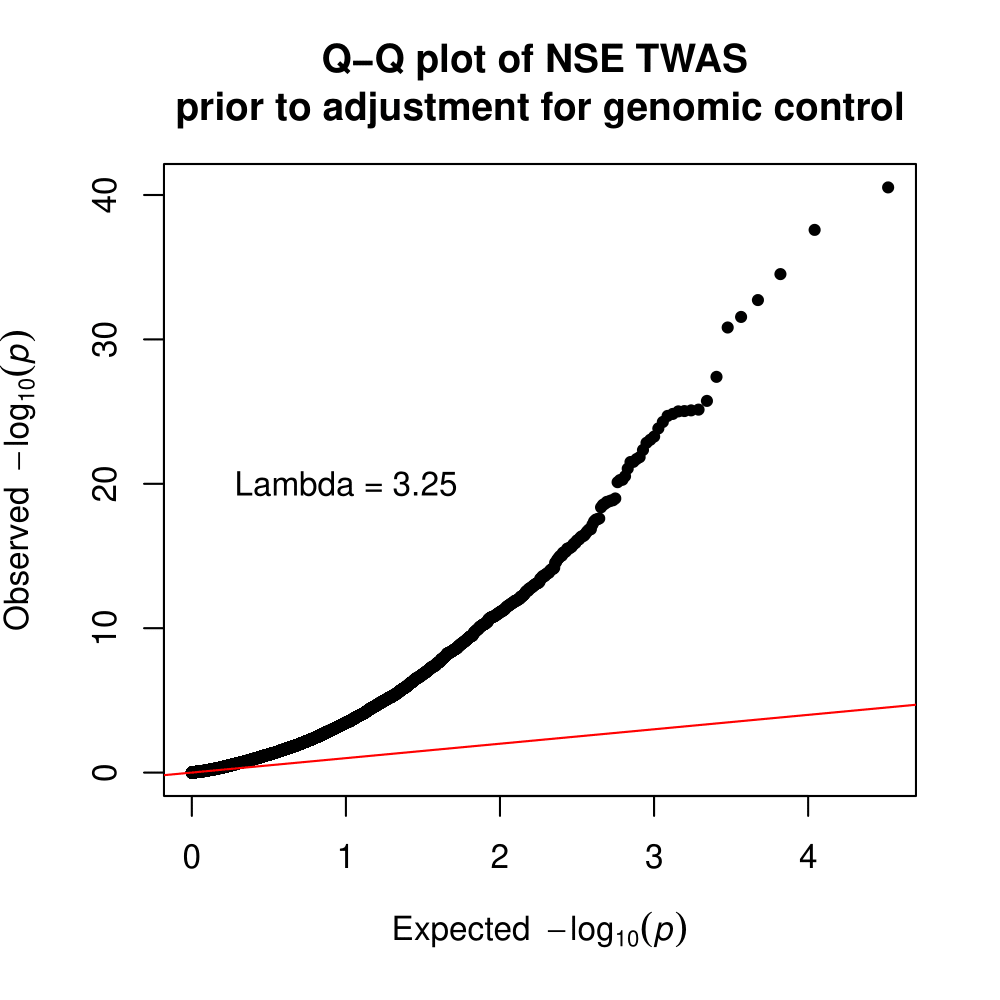

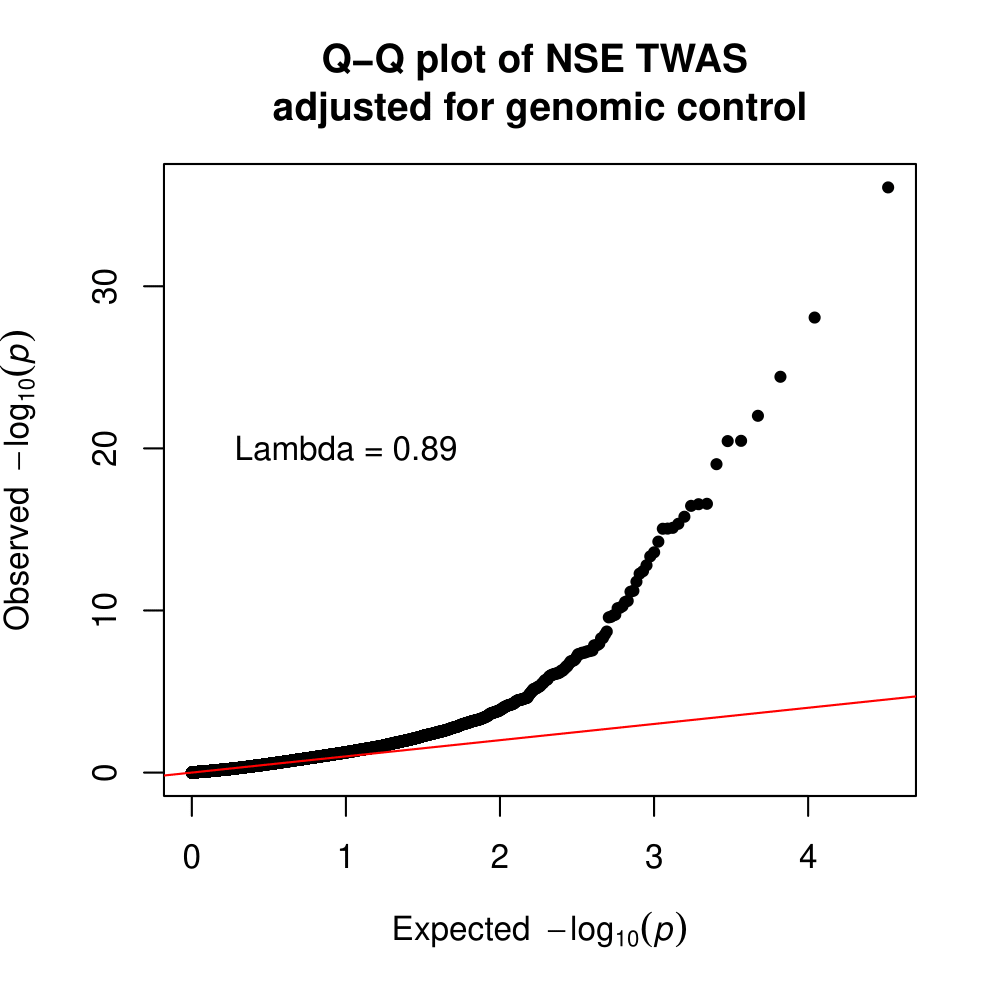

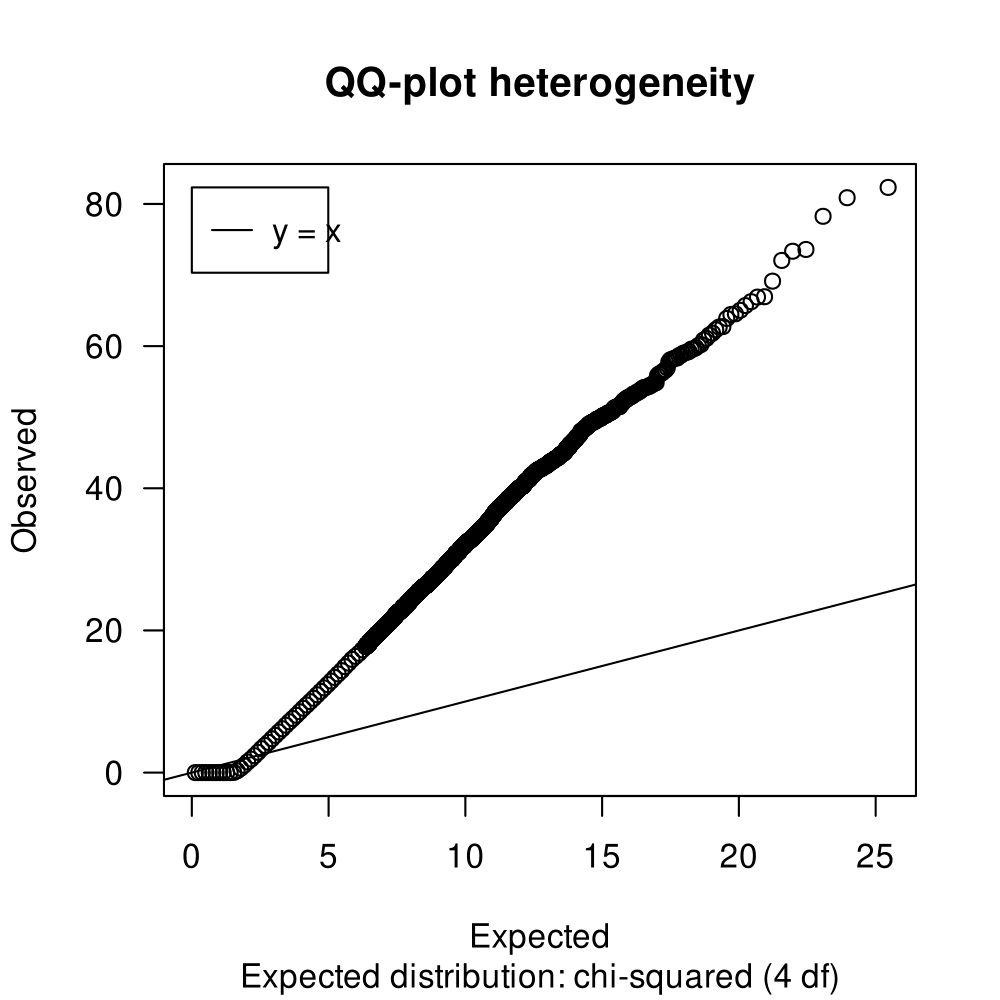
Fig S4. QQ plot of the Cochran’s test for the transcriptomic meta-analysis.

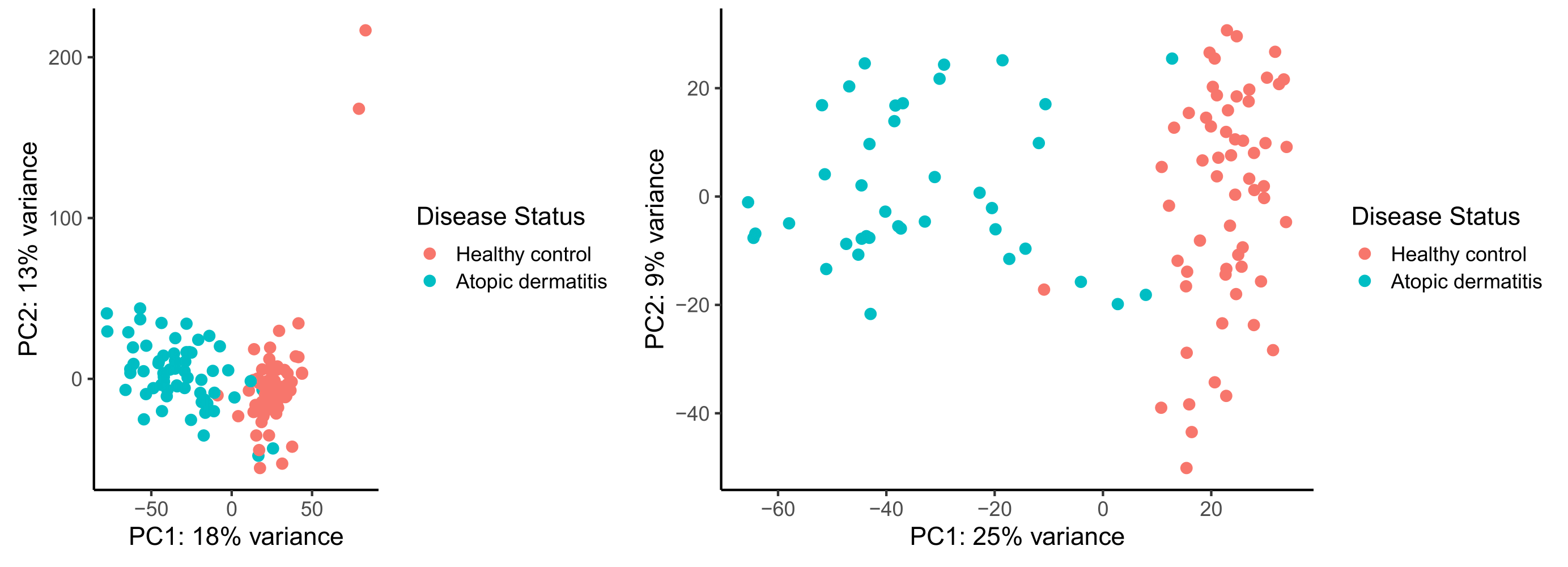
Fig S5. Principal component analysis (PCA) pre (left) and post (right) outlier correction based on the ROBPCA algorithm.

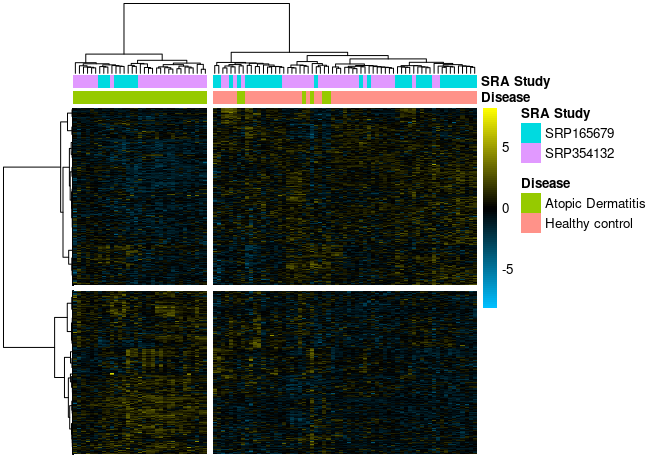
Fig S6. Heatmap of transcripts clustered during the WGCNA analysis. Transcripts falling in the gray module (ME0) were excluded from the heatmap.

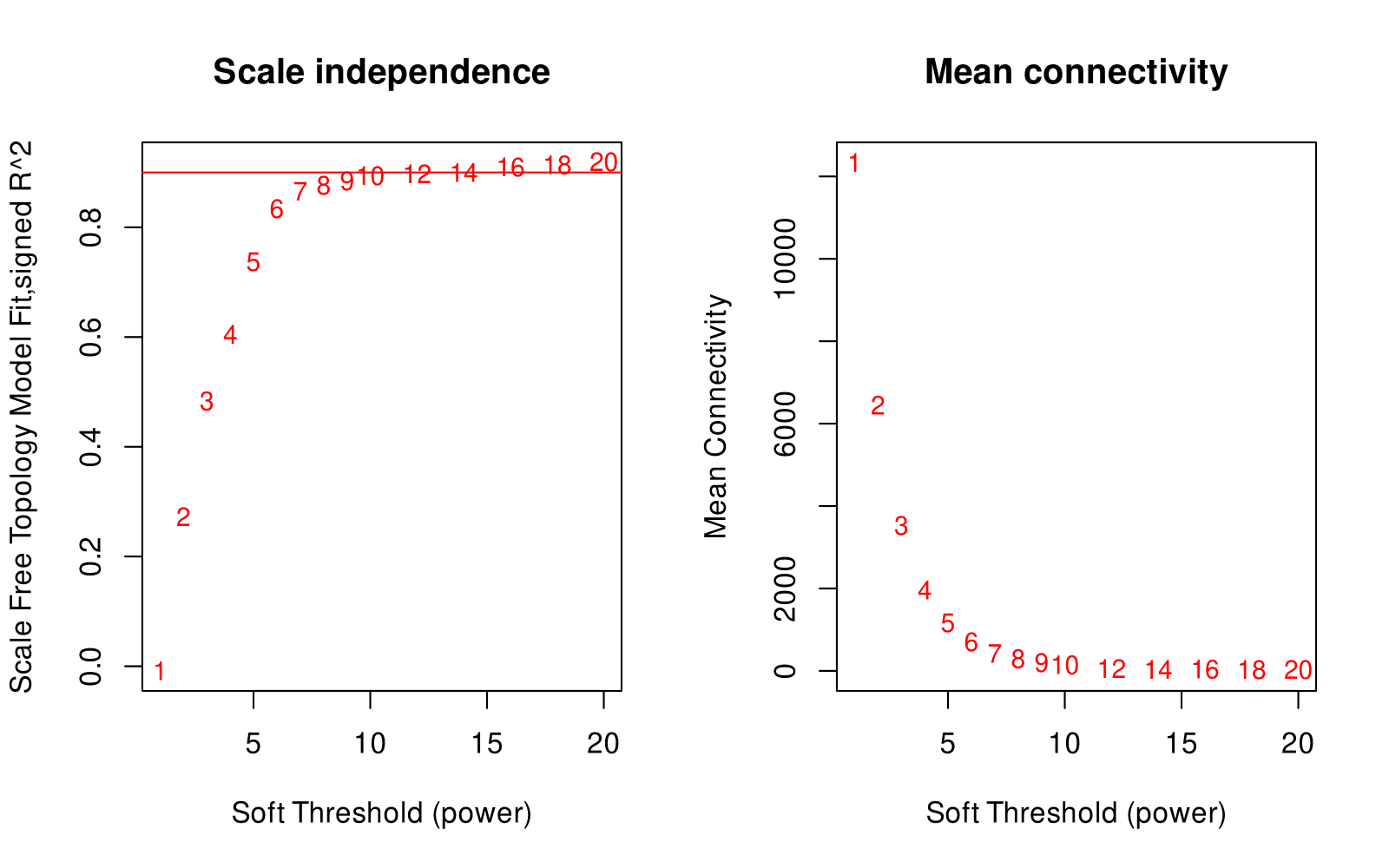
Fig S7. Determination of the soft-thresholding power for WGCNA.

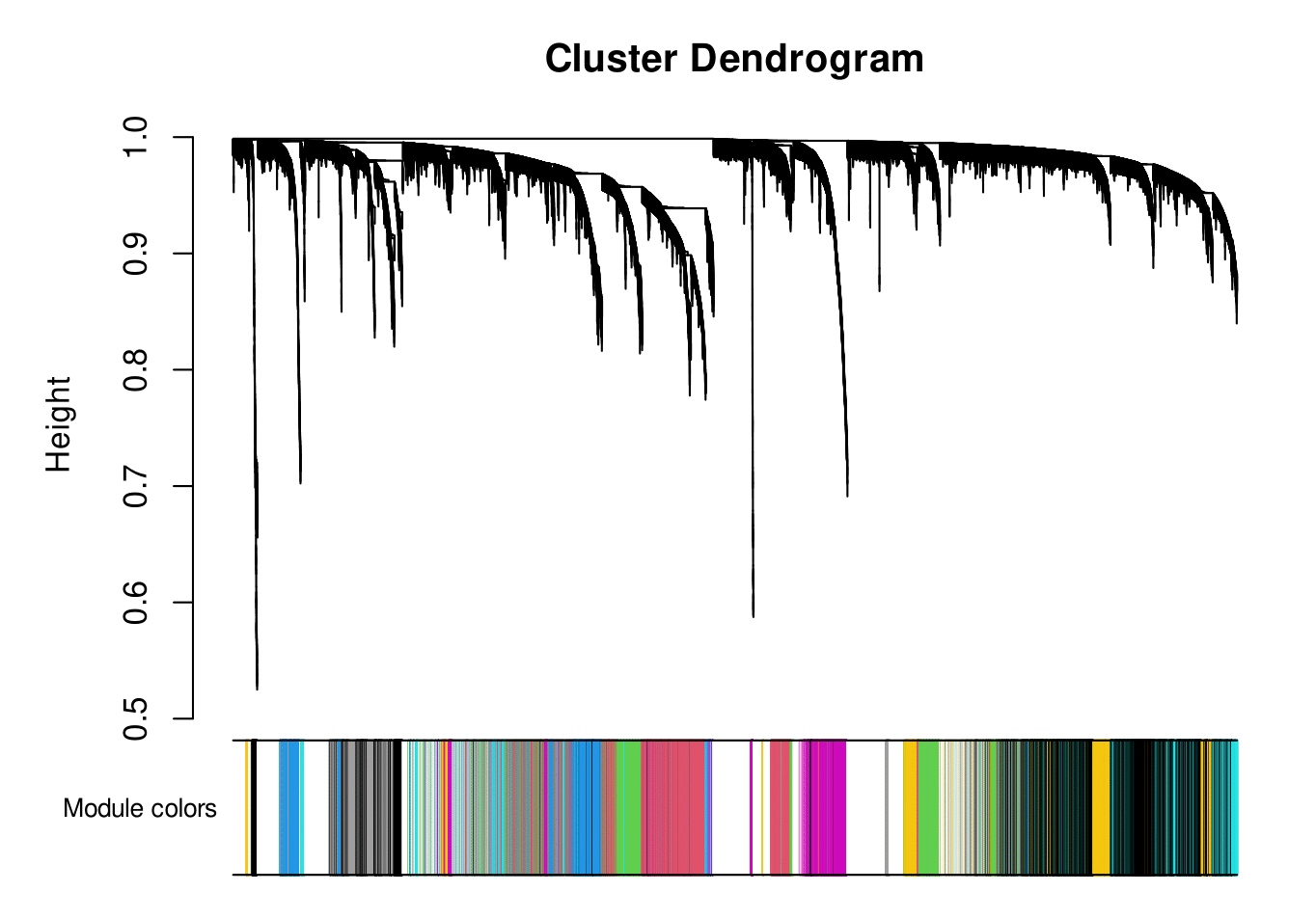
Fig. S8. Cluster dendrogram and colored co-expression modules.
